# Estimating COVID-19 Virus Prevalence from Records of Testing Rate and Test Positivity

**DOI:** 10.1101/2020.11.17.20233643

**Authors:** Arnout JW Everts

## Abstract

**Introduction:** PCR testing for COVID-19 is not done at random but selectively on suspected cases. This paper presents a method to estimate a “genuine Virus Prevalence” by quantifying and removing the bias related to selective testing.

**Methods:** Data used are from nine (9) neighbouring countries in Western Europe that record similar epidemic trends despite differences in Testing Rate. Regression analysis is used to establish a relationship of declining Test Positivity with increased Testing Rate. By extrapolating this trend to an “infinitely complete” Testing Rate, an unbiased Test Positivity or “genuine Virus Prevalence” is computed. Via pairing of “genuine Virus Prevalence” with Excess-Deaths, a “genuine Infection Fatality Rate (IFR) is also derived.

**Results:** Peak levels of “genuine Virus Prevalence” were around 0.5 to 2% during the 1^st^ epidemic “wave” (week 10 to week 20) and are approaching similar levels in the ongoing 2^nd^ “wave” (week 34 onward). “Genuine Virus Prevalence” estimates are close to reported Seroprevalence in the studied countries with a correlation coefficient of 0.58. “Genuine” IFR is found comparable to closed-community model IFR. Finally, results of community mass-testing in Slovakia are within the estimated range of “genuine Virus Prevalence”.

**Conclusions:** Estimates of “genuine Virus Prevalence” benchmark favourably to other indications of virus prevalence suggesting the estimation method is robust and potentially deployable beyond this initial dataset of countries. “Genuine Virus Prevalence” curves suggest that during the 1^st^ epidemic “wave”, curve flattening and waning happened at very modest levels of infection spread, either naturally or facilitated by government measures.

## 1. Introduction

According to the World Health Organization (WHO), Corona virus disease 2019 (COVID-19) reached the pandemic phase on March 11, 2020. As of October 22, 2020, it had spread to more than 200 countries worldwide, leading to 41,333,175 registered infections and 1,132,869 deaths. As the pandemic evolved, countries implemented routine testing of suspected cases using real-time RT-PCR test assays; initially for diagnostic and reporting purpose but subsequently stepping-up the Testing Rate in an attempt to map community spread of the virus and to guide containment measures like isolation of infected patients, lockdown of neighbourhoods with a high number of reported cases, etc. Obviously the testing was not deployed at random but primarily on suspected cases especially early in the epidemic when test capacity was very limited. Because of this biased deployment of testing, the reported percentage of positive tests (the so-called “Test Positivity”) is almost certainly higher than the true percentage of infected members of the population (called “genuine Virus Prevalence” throughout this paper). It is likely that as countries stepped up the testing beyond a baseload of symptomatic patients to include groups with a likely lower virus prevalence (mild cases, routine testing of e.g. travellers and sportsmen, etc.), this biasing effect reduced. In other words, the biasing of Test Positivity due to biased testing may have reduced as Testing Rate increased. A study of testing records from the US, relatively early in the epidemic outbreak, already demonstrated and quantified some of this effect with a sample-selection model [1]. This paper attempts to quantify and remove sampling bias from reported test results using a much larger European dataset covering a longer period, using regression statistics similar to methods deployed before on Malaria survey data [2]. Estimates of “genuine Virus Prevalence” are made by analysing the relationship between Testing Rate (number of tests per week per capita) and Test Positivity over time, for a number of countries within the same geographic realm and therefore subject to similar trends in virus prevalence.

## 2. Methods

This paper made use of open-domain data on COVID-19 reported Cases, Deaths, Testing Rate and Test Positivity per country per week as published by the European Centre for Disease Prevention and Control (ECDC [3]). COVID-19 records available from this database up to 16/10/2020 were used.

Nine (9) countries in Western Europe were selected for analysis considering neighbouring geography and consistency in reporting of data especially of Testing Rate and Test Positivity. The idea being that epidemic trajectories across these neighbouring countries might be sufficiently similar to observe the impact of Testing Rate on reported Test Positivity, as the testing rate in different countries was stepped up over time but with different pace and timing. And to then quantify this impact through statistical regression analysis. A trend between Test Positivity and Testing Rate was established and then used to correct individual Test Positivity records (reports of Test Positivity per week per country) for the anticipated testing bias. This correction involves extrapolation of the trend between Testing Rate and Test Positivity to an “infinitely complete” Testing Rate of 100,000 tests per 100,000 people per week. Bias-corrected records of Test Positivity are considered an estimate of what Test Positivity might have been if testing were complete, in other words, how the genuine infection spread among the community might have varied over time in the different countries. In this paper, this entity is called “genuine Virus Prevalence”, calculated as follows:

> genuine Virus Prevalence = reported Test Positivity / regressed_Positivity [TR_a_] * regressed_Positivity [TR_100,000_]

Where:

> *regressed_Positivity* refers to the relationship between Test Positivity and Testing Rate, TR_a_ being the actual Testing Rate reported for each country Test Positivity record and TR_100,000_ being an “infinitely complete” testing rate of 100,000 tests per 100,000 persons per week.

In the above equation, *regressed_Positivity* [TR_*100,000*_] is shifted up (P10) or down (P90) by 1.28 times the regression *Standard Error* to define a confidence band around the best-fit estimates of “genuine Virus Prevalence”.

Estimated “genuine Virus Prevalence” per week and per country is considered indicative of the number of newly detected infections in that week rather than the total number of active infections (new plus existing still active infections). Reason being, the weekly PCR Test Positivity data from which “genuine Virus Prevalence” is derived, presumably record only the new active infections confirmed for each week (CDC recommends not to subject cases with a confirmed positive test result to another PCR for some three months following recovery [4]).

To validate the estimates of “genuine Virus Prevalence”, they were compared against available estimates of COVID-19 Seroprevalence for the studied countries, mostly from blood plasma studies [5, 6, 7, 8, 9, 10, 11, 12]. The difference in reporting period needs to be taken into consideration. Whilst “genuine Virus Prevalence” is like an “instantaneous” measure of virus prevalence as it is based on active infections seen by PCR testing, Seroprevalence data reflects antibody response to either active or historic infections that may date back several months [13]. In this paper, it is assumed that the average antibody residence time is about three months (12 weeks). Therefore, for each Seroprevalence report the cumulative “genuine Virus Prevalence” over the reporting period plus the 12 weeks preceding it, is reflected against the Seroprevalence percentage.

This study also paired estimates of “genuine Virus Prevalence” per week per country, with estimates of COVID-19 fatality per week per country, to yield a “genuine” Infection Fatality Rate (IFR). In view of the general belief that officially recorded numbers of confirmed COVID-19 Deaths likely underestimate the true fatality impact of COVID-19 due to a variety of diagnosis and reporting issues [14], [15], Excess Deaths per country per week were used instead. Excess Deaths were extracted for the 1^st^ epidemic “wave” only (defined in this instance as the period from 15th March to 7^th^ July 2020) to avoid any bias due to incompleteness of Excess Deaths data for recent weeks, and compared to “genuine virus prevalence” for the same period. Source of data was the Our World In Data website [15] and data was downloaded as of 30/10/2020. “Genuine” IFR values were then benchmarked against IFR estimates from a closed-community outbreak study: the Diamond Princess (DP) cruise ship [16]. Because of the strong age-dependency of IFR shown by the DP study [16] and by the confirmed COVID-19 death statistics broken down by age, IFR estimates were corrected for the differences in age breakdown between the DP closed-community and each individual European country, using demographics data obtained from the UNdata website [17] (downloaded as of 16/10/2020). This comparison of “genuine IFR” against closed-community IFR comprises a second attempt of results validation.

A final benchmarking of “genuine Virus Prevalence” estimates is done against results of mass-testing of the Slovakia population at end of Week 43 [18]. To facilitate this benchmarking, Testing Rate and Test Positivity data for that week were obtained from the Slovak Municipal health department [19], complementing the records of earlier weeks obtained from ECDC [3].

## 3. Results

### 3.1. Estimating Genuine Virus Prevalence

All of the nine European countries record a similar two “waves” epidemic trajectory comprising a 1^st^ “wave” in week 10 to week 20 and a 2^nd^ “wave” from about week 34 onward. This is evident in both the number of reported Cases as well as in the % of Positive test outcomes (“Test Positivity Rate”). Similarity in epidemic trends is despite the Testing Rate (number of tests per week per capita) being quite different from one country to another. For example, Portugal roughly doubled its Testing Rate from about 690 tests per 100,000 population per week around the peak of the 1^st^ epidemic “wave” (week 15-16), to 1,440 in the ongoing 2^nd^ “wave” (week 40). Whilst over the same period, United Kingdom stepped up its testing more than ten-fold: from 150 test per 100,000 population per week in week 15-16, to 2,710 in week 40. Differences in Testing Rate over time and between countries are likely to reflect a varying degree of test selectivity bias on recorded Positivity.

A weighted average of Test Positivity across all countries was computed and used to quantify the differences in Test Positivity in different countries, over time and relative to cross-country trend. Differences in Test Positivity between countries may reflect a variety of things including genuine differences in the spread of COVID-19 among the population, testing protocol, etc etc., but certainly inclusive of differences in test-selectivity “bias” due to different Testing Rate. To demonstrate this, Figure 1 cross-plotted the difference between individual-country Test Positivity and the cross-country-average Test Positivity, against Testing Rate. Note that the difference is expressed as a ratio (in other words, the Y-axis of Figure 1 shows *individual-country Test Positivity* over *cross-country-average Test Positivity*). As expected, there is a clear trend of decreasing Test Positivity with increased Testing Rate. A best-fit relationship was computed through regression analysis as shown in Figure 1. Given there are many other factors that may influence the difference in test Positivity between countries and hence it cannot be expected that all differences are explained by Testing Rate, the author considers the observed Correlation Coefficient (R^2^) of 0.215 as statistically significant.

**Figure 1:**
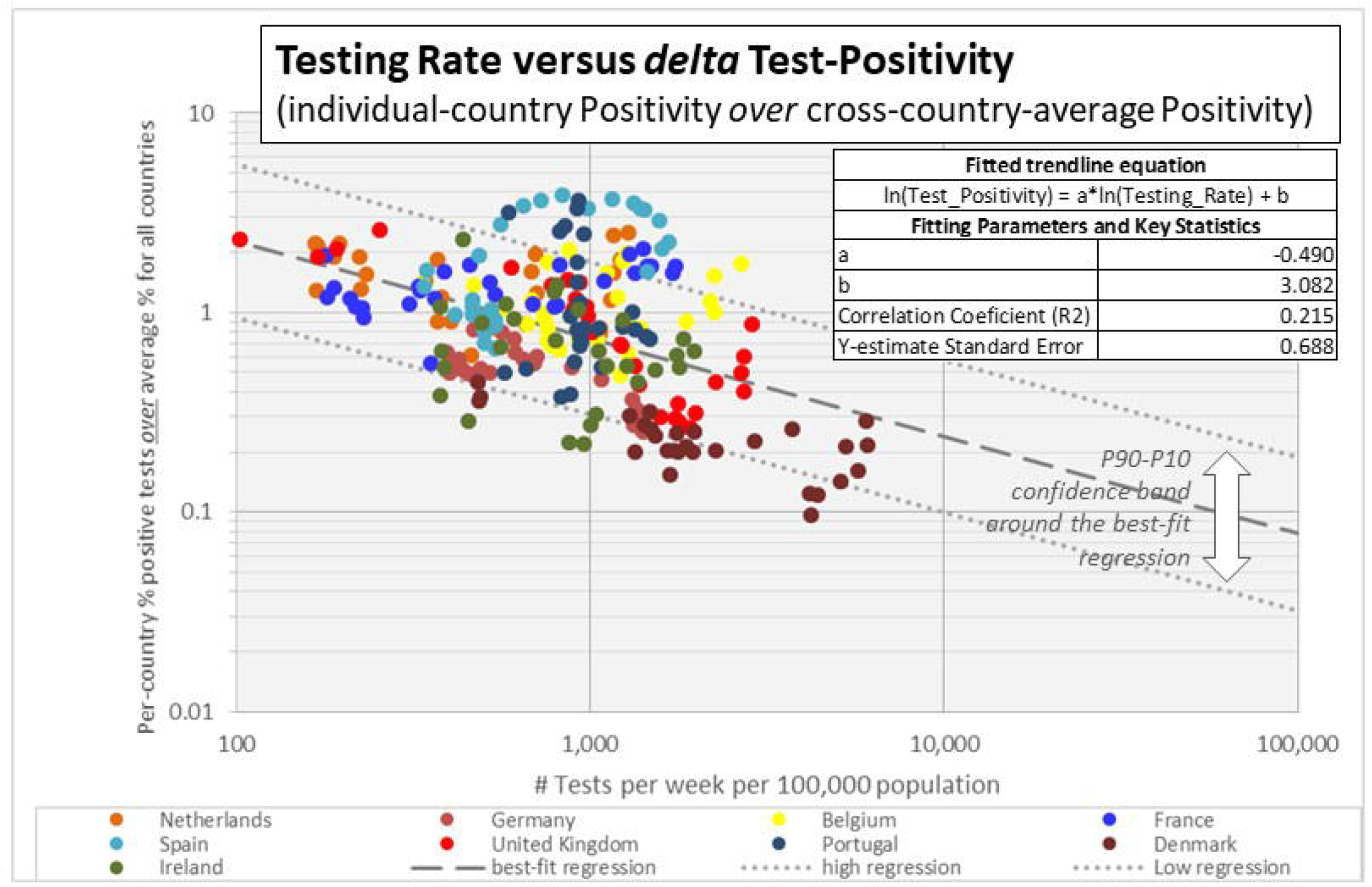
Testing Rate versus delta-Test Positivity for the nine European countries studied.

Figure 2 shows trajectories of the best-estimate of “genuine Virus Prevalence”: Test Positivity corrected for suspected test-selectivity bias. As explained in the Methods section of this paper, correction was done by extrapolating the regressed trend between Testing Rate and Test Positivity (Figure 1) to an infinitely complete Test Rate of 100,000 tests per 100,000 population per week. Peak levels of “genuine Virus Prevalence” were around 0.5 to 2% during the 1^st^ epidemic “wave”, depending on the country, and are approaching similar levels in the ongoing 2^nd^ epidemic “wave”. Obviously these values are per-country averages and it is possible that at individual epidemic-outbreak localities, the true virus prevalence might have been considerably higher. The author believes that “genuine Virus Prevalence” may be a fair reflection of the degree of community spread of COVID-19 infections. Next paragraphs will validate this hypothesis by comparing “genuine Virus Prevalence” with independent indicators of prevalence.

**Figure 2:**
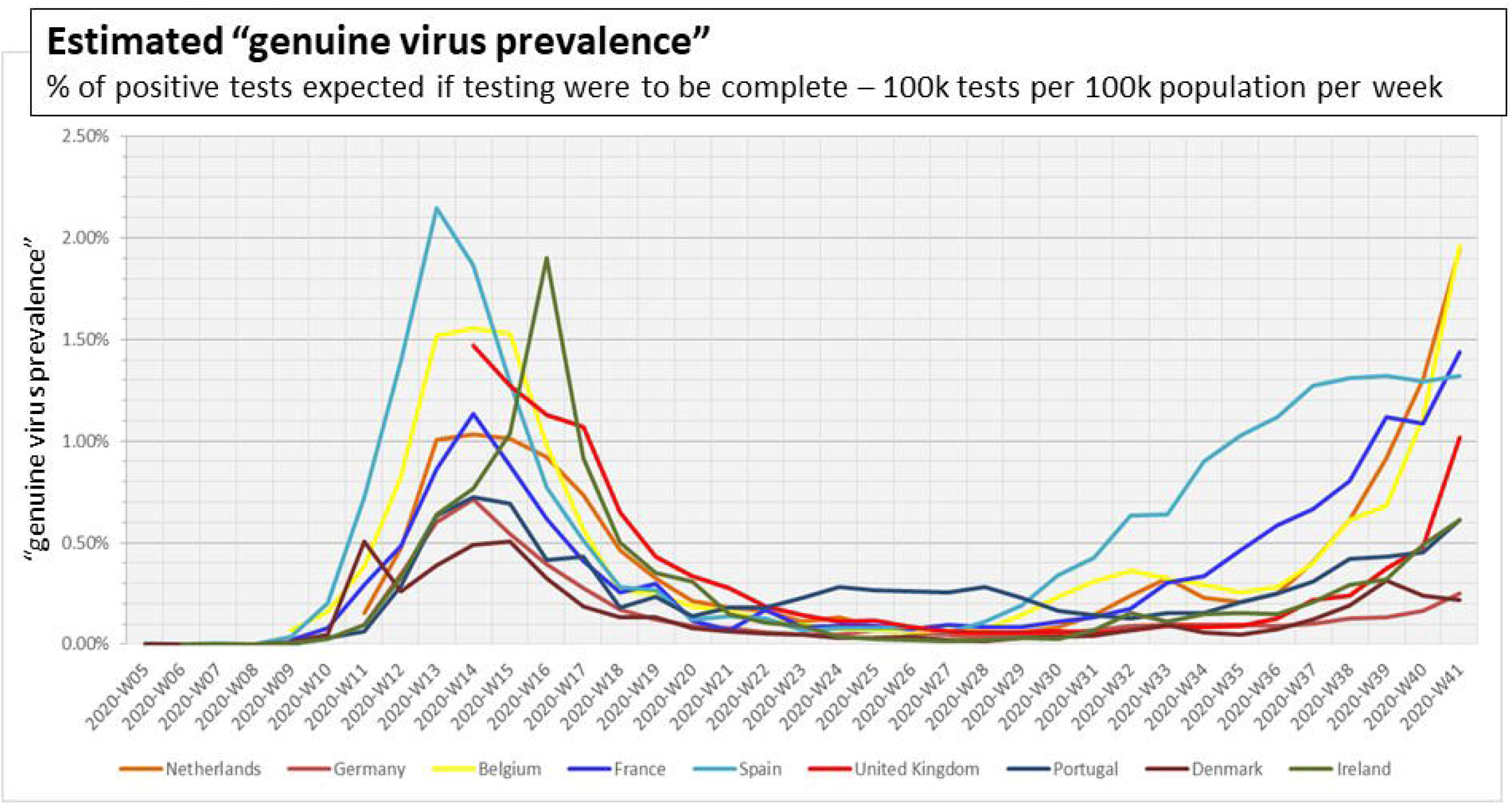
“genuine Virus Prevalence” over time for the nine European countries studied.

### 3.2. Genuine Virus Prevalence versus Seroprevalence

Table 1 shows a benchmarking of “genuine Virus Prevalence” estimates against Seroprevalence data available for the studied countries. As explained, reported Seroprevalence reflects antibody response to active plus historic infections over a considerable time period (this paper assumed 12 weeks on average), Therefore, Table 1 compares Seroprevalence percentages against cumulative “genuine Virus Prevalence” over the 12 weeks preceding the Seroprevalence reporting date. Seroprevalence is generally somewhat lower than the equivalent best estimates of “genuine Virus Prevalence” but the difference is not much. 86% of the Seroprevalence data fall within the P90-P10 confidence band of “genuine Virus Prevalence”. The correlation coefficient (R^2^) between Seroprevalence and cumulative “genuine Virus Prevalence” is 0.58. Figure 4 illustrates the degree of correspondence between these two independent measures of community virus-spread.

**Table 1:** Comparison of available Seroprevalence data against estimated ‘genuine Virus Prevalence” for the studied countries

| Country | Reporting Period | Sero-prevalence | Genuine Virus Prevalence |  |  | Source / Remark |
| --- | --- | --- | --- | --- | --- | --- |
|  |  |  | P90 | Best | P10 |  |
| Netherlands | 11/5 - 18/5 | 6.5% | 2.6% | 6.3% | 15.3% | antibody testing on blood plasma samples [5] |
| Germany | April - June | 1.3% | 1.5% | 3.6% | 8.8% | Tests on nearly 12,000 German blood donors [6] |
| Belgium | 30/3 - 5/4 | 2.9% | 1.2% | 2.9% | 7.1% | Antwerp University antibody study in blood serum [7] |
|  | 20 - 26/4 | 6.0% | 2.9% | 7.0% | 16.9% |  |
|  | 18 - 25/5 | 6.9% | 3.4% | 8.3% | 20.0% |  |
|  | 8 - 13/6 | 5.5% | 3.3% | 8.0% | 19.4% |  |
|  | 29/6 - 5/7 | 4.5% | 1.8% | 4.4% | 10.5% |  |
| France | W12-W13 | 2.7% | 0.7% | 1.7% | 4.2% | blood donor study [8] |
| Spain | 27/4 - 11/5 | 5.0% | 2.6% | 6.4% | 15.4% | nationwide seroepidemiological study, point of care test [9] |
|  |  | 4.6% | 2.6% | 6.4% | 15.4% | nationwide seroepidemiological study, immunoassay [9] |
| United Kingdom | 6-29/5 | 8.3% | 2.7% | 6.6% | 16.0% | seroprevalence in blood donors [10] |
|  | 19/8-13/9 | 6.1% | 0.6% | 1.4% | 3.4% | seroprevalence in blood donors [10] |
| Portugal | 21/5-8/7 | 2.9% | 2.3% | 5.5% | 13.3% | first Covid-19 National Serological Survey Portugal [11] |
| Denmark | 22/6 -10/8 | 2.8% | 0.9% | 2.3% | 5.4% | blood antibodies study of Falck employees [12] |

**Figure 3:**
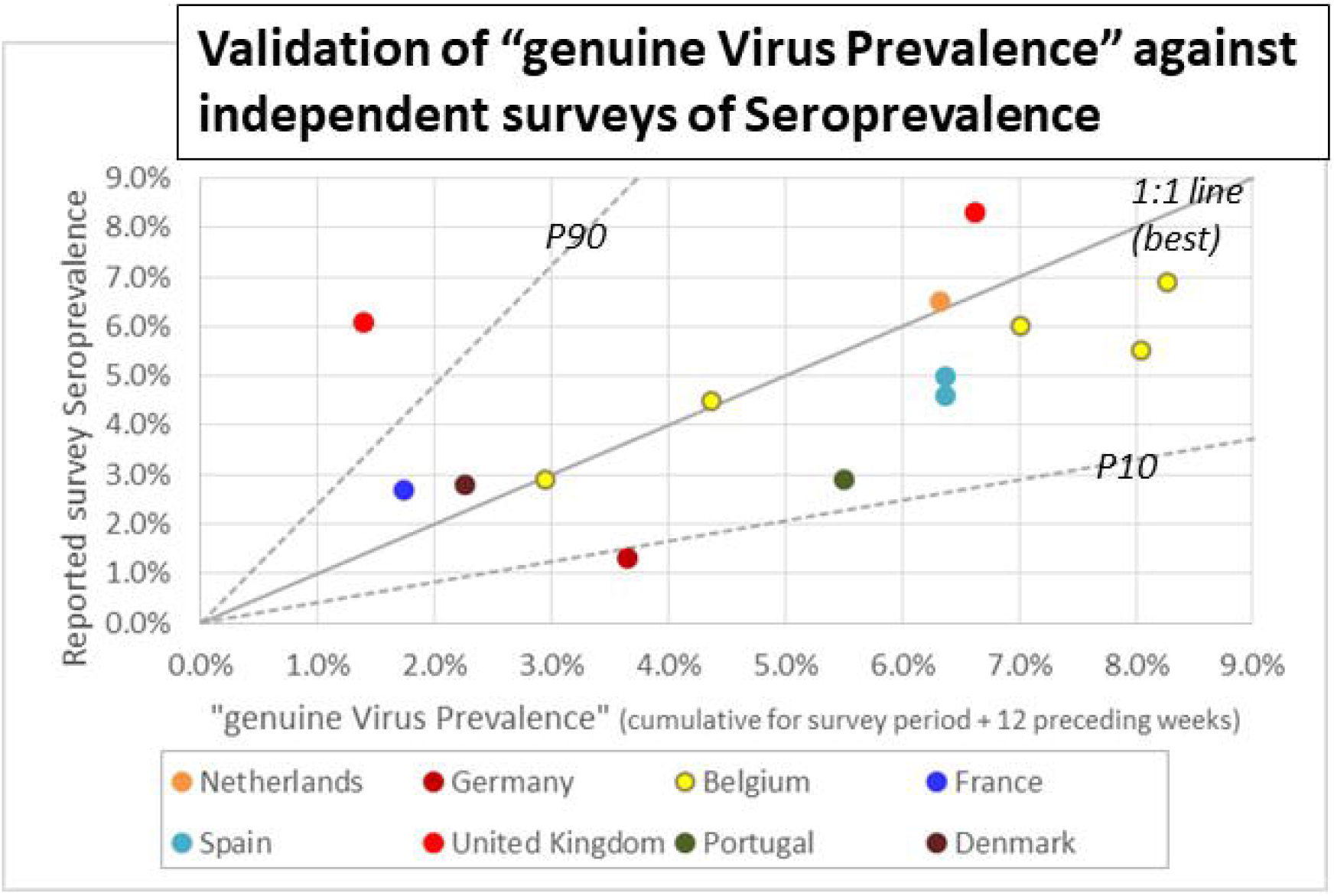
Cross-plot of reported Seroprevalence against the cumulative of “genuine Virus Prevalence” over the survey period + 12 preceding weeks (for rationale see text). The P10-P90 confidence band around the best estimate of “genuine Virus Prevalence” is indicated

**Figure 4:**
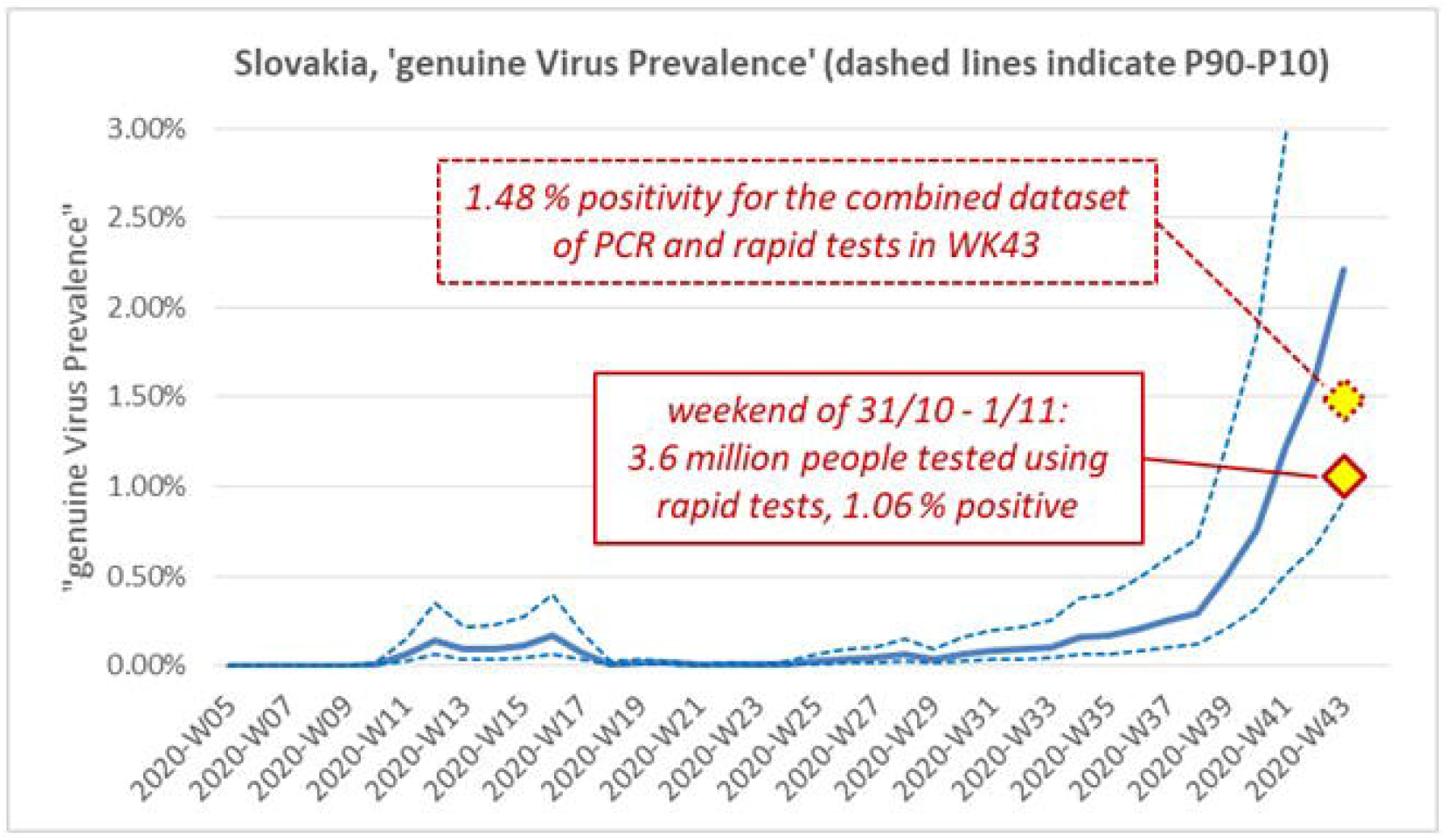
“genuine Virus Prevalence” estimated for Slovakia, compared against results of recent community mass-testing using rapid antigen method.

### 3.3. Genuine Infection Fatality Rate

Estimates of “genuine Virus Prevalence” (which are, as explained in the Methods section, believed to reflect the number of new active infections reported in a week) were converted to cumulative and then paired with estimates of Excess Deaths per week and per country, to yield “genuine” IFR estimates (Table 2). As explained, Excess Deaths were extracted and used to estimate IFR for the period 15^th^ March to 7^th^ July only (the 1^st^ epidemic “wave”). Note that no Excess Deaths data was available for Ireland. With the exception of Belgium and Denmark, Excess Death numbers are higher than the confirmed COVID-19 Deaths and in some instances, considerably higher (Table 2).

**Table 2:**
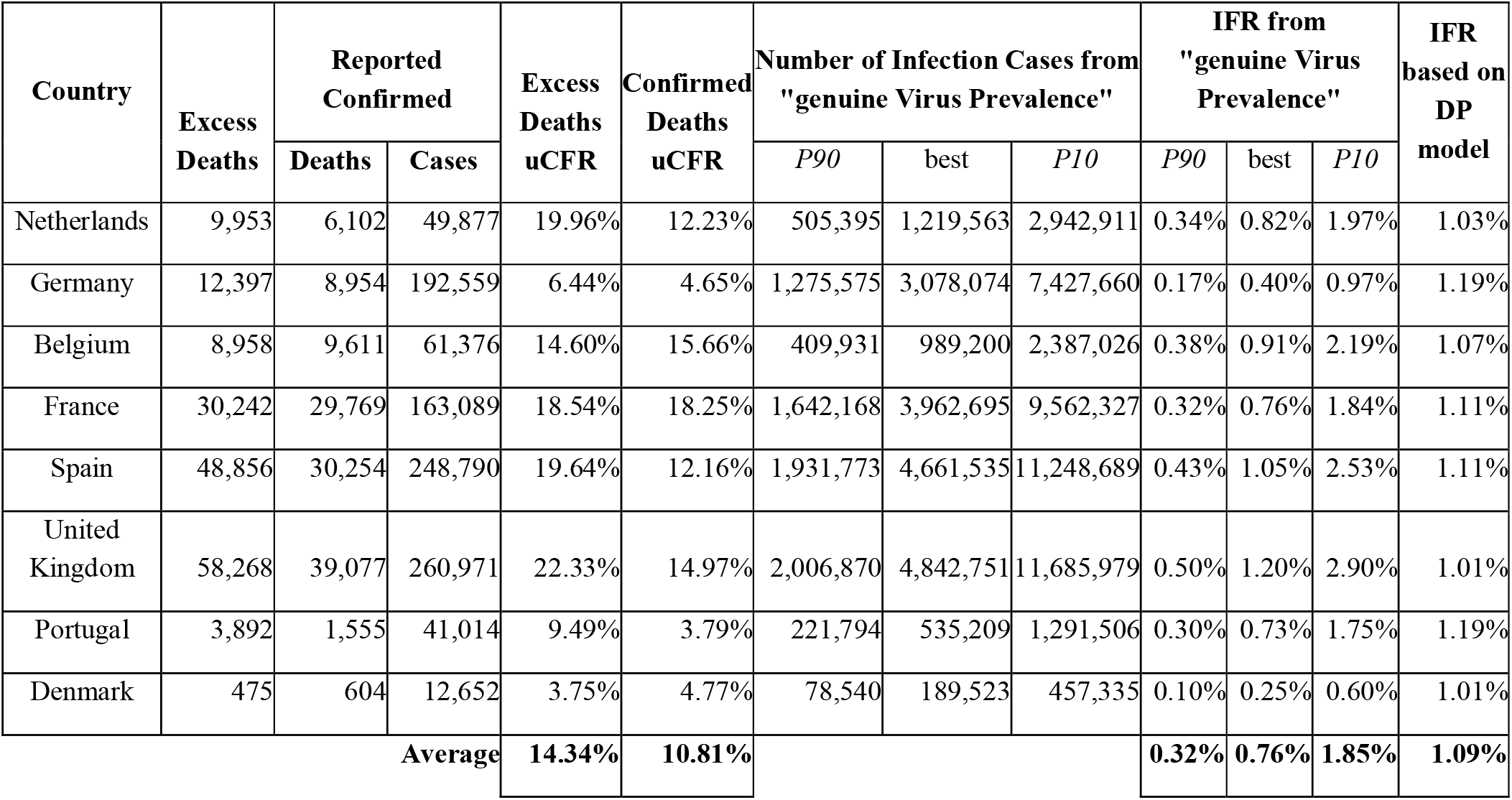
Excess deaths, Confirmed Cases and Deaths, uncorrected Case Fatality Rates (uCFR), number of Infection cases from “genuine Virus Prevalence and corresponding Infection Fatality Rate (IFR) estimates compared against IFR from closed-community data (DP cruise ship [14]). Estimates are for the period 15th March to 7th July only (the 1st epidemic “wave”) except for UK where the analysis starting-date is 5th April (start of test-data records).

| Country | Excess Deaths | Reported Confirmed |  | Excess Deaths uCFR | Confirmed Deaths uCFR | Number of Infection Cases from "genuine Virus Prevalence" |  |  | IFR from "genuine Virus Prevalence" |  |  | IFR based on DP model |
| --- | --- | --- | --- | --- | --- | --- | --- | --- | --- | --- | --- | --- |
|  |  | Deaths | Cases |  |  | P90 | best | P10 | P90 | best | P10 |  |
| Netherlands | 9,953 | 6,102 | 49,877 | 19.96% | 12.23% | 505,395 | 1,219,563 | 2,942,911 | 0.34% | 0.82% | 1.97% | 1.03% |
| Germany | 12,397 | 8,954 | 192,559 | 6.44% | 4.65% | 1,275,575 | 3,078,074 | 7,427,660 | 0.17% | 0.40% | 0.97% | 1.19% |
| Belgium | 8,958 | 9,611 | 61,376 | 14.60% | 15.66% | 409,931 | 989,200 | 2,387,026 | 0.38% | 0.91% | 2.19% | 1.07% |
| France | 30,242 | 29,769 | 163,089 | 18.54% | 18.25% | 1,642,168 | 3,962,695 | 9,562,327 | 0.32% | 0.76% | 1.84% | 1.11% |
| Spain | 48,856 | 30,254 | 248,790 | 19.64% | 12.16% | 1,931,773 | 4,661,535 | 11,248,689 | 0.43% | 1.05% | 2.53% | 1.11% |
| United Kingdom | 58,268 | 39,077 | 260,971 | 22.33% | 14.97% | 2,006,870 | 4,842,751 | 11,685,979 | 0.50% | 1.20% | 2.90% | 1.01% |
| Portugal | 3,892 | 1,555 | 41,014 | 9.49% | 3.79% | 221,794 | 535,209 | 1,291,506 | 0.30% | 0.73% | 1.75% | 1.19% |
| Denmark | 475 | 604 | 12,652 | 3.75% | 4.77% | 78,540 | 189,523 | 457,335 | 0.10% | 0.25% | 0.60% | 1.01% |
| Average |  |  |  | 14.34% | 10.81% |  |  |  | 0.32% | 0.76% | 1.85% | 1.09% |

Compared to uncorrected Case Fatality Rate (uCFR) estimates derived from pairing the cumulative number of confirmed Cases with Excess Deaths per country, “genuine” IFR estimates are between 13 to 25 times lower. Most of the “best” estimates of “genuine IFR” are relatively close to closed-community model IFR [16] (Table 2). 75% of the model-IFR estimates fall within the P90-P10 confidence band of “genuine IFR”.

### 3.4. Genuine Virus Prevalence versus Slovakia Mass Rapid-Testing

Slovakia recently became the first country to conduct mass testing of its population to estimate the scale of COVID-19 community spread. A total of 3,625,322 Slovaks over the age of nine were given antigen swab tests over the weekend of 31/10 to 1/11; 38,359 (1.06%) tested positive [18]. In addition, during the same week as the mass rapid testing Slovakia also conducted a total of 112,952 PCR tests of which 14.8% positive. The total number of newly detected cases for week 43 is therefore 55,033 on 3,738,274 tests (1.48% Positivity).

Slovakia is not included in the nine countries to which this paper fitted the equations for estimating “genuine Virus Prevalence” from records of Test Positivity and Testing Rate, but it is geographically near the analysed countries and recording similar epidemic trends. Therefore, this paper deployed the same methodology as used for the other countries to compute a “Genuine Virus Prevalence” for Slovakia. Figure 4 shows the “genuine Virus Prevalence” curve against the community-scale rapid testing result. The range in “genuine Virus Prevalence” (from bias-corrected PCR Test Positivity only) estimated for Slovakia in Week 43 is 0.92% / 2.21% / 5.34% (P90 / Best / P10) which is reasonably close to the mass testing findings.

## 4. Discussion

### 4.1. Validation of “genuine Virus Prevalence” estimates

This paper presented a relatively simple and straightforward method of estimating “genuine Virus Prevalence” based on records of Test Positivity and Testing Rate over time. Three attempts of results validation were presented namely 1) by comparison of “genuine Virus Prevalence” against Seroprevalence data, 2) via estimation of “genuine IFR” which was then cross-compared against closed-community IFR, and 3) by estimating “genuine Virus Prevalence” for Slovakia and then comparing the outcome with results of the recent community mass-testing in that country. Paragraphs below discuss whether the validation results can be considered favourable.

#### 4.1.1. Validation against Seroprevalence

Differences between Seroprevalence for certain countries and cumulative “genuine Virus Prevalence” in the 12 weeks prior to Seroprevalence reporting, are generally quite small (Table 1) and the R2 of 0.58 is suggestive of a significant correlation (Figure 4). The fact that “genuine prevalence” is generally somewhat higher than reported Seroprevalence is unsurprising. Seroprevalence studies are mostly using blood-bank data that is likely biased towards asymptomatic COVID-19 cases. Symptomatic patients would not be allowed as blood donors during or shortly after suffering from the disease.

Uncertainty on the exact duration of test-detectable antibody response following an infection may also explain some of the observed discrepancy between Seroprevalence and equivalent estimates of “genuine Virus Prevalence”. If in reality the antibody count in some patients wanes to undetectable in less than 12 weeks, it means that for a fair comparison “genuine Virus Prevalence” should be summed over a period of less than 12 weeks and/or with a tapering window. In any case, such would lower the “genuine Virus Prevalence” numbers and bring them even closer to Seroprevalence.

All in all, the comparison between “genuine Virus Prevalence” and Seroprevalence is considered favourable.

#### 4.1.2. Validation via Infection Fatality Rate

“Genuine” IFR values computed from the pairing of “genuine Virus Prevalence” with Excess Deaths are reasonably close to closed-community model IFR (Table 2), albeit slightly lower. The author identified at least two factors that can explain why “genuine” IFR estimates are on the low side compared to model IFR. Firstly, given the nature of the underlying PCR Test Positivity this paper assumes that “genuine Virus Prevalence” (per week, per country) is reflective of the newly discovered COVID-19 cases in that week. If this assumption is not entirely correct and in reality, “genuine Virus Prevalence” reflects a mix of new and existing cases, it means that the true number of new cases per week is less than the “genuine Virus Prevalence”. Such would reduce the cumulative number of cases and consequently, increase the IFR. A second, alternative explanation could be that the closed-community IFR models are based on observations early in the pandemic. Since then, insights in the disease and effectiveness of treatment methods have evolved. It is entirely possible that some of this progress is reflected in a genuine drop in IFR from the current data compared to early-stage datasets like the Diamond Princess cruise-ship.

Given these considerations and interpretation uncertainties, this author believes that the match between “genuine” IFR and closed-community model IFR is close enough to conclude that the “genuine” IFR estimates and the underlying “genuine Virus Prevalence” are reasonable.

#### 4.1.3. Validation against Slovakia Mass Testing

Test Positivity of 1.06% as recorded in the COVID-19 mass community-testing in Slovakia, week falls towards the low end of the range of “genuine Virus Prevalence” estimated for that week. (Figure 4). Positivity for the combined test dataset of rapid tests and PCR tests during that week (1.48%) is closer to the “genuine Virus Prevalence” best estimate but still on the low side. This observation is consistent with the concern expressed by some experts [20], [21] that antigen rapid-tests are less reliable and possibly much less sensitive compared to PCR tests.

Slovakia was not part of the initial dataset of nine European countries from which this paper derived the method and correction factors for estimating “genuine Virus Prevalence” from Test Positivity and Testing Rate. Nevertheless, “genuine Virus Prevalence” for Slovakia is in the same range as the community spread suggested by mass testing. This observation suggests that the correction method and correction factors proposed herein may be more universally deployable, beyond the initial dataset of countries.

### 4.2. Significance of the trends in “genuine Viral Prevalence”

The “genuine Viral Prevalence” curves for the studied European countries consistently show a two “waves” pattern (Figure 1) with a 1^st^ epidemic “wave” in week 10 to week 20 and a 2^nd^ “wave” from about week 34 onward. “Genuine Virus Prevalence” for the 2^nd^ “wave” to date is approaching similar levels as during the peak of the 1^st^ “wave”. This means that the higher number of confirmed COVID-19 Cases in the ongoing 2^nd^ “wave” (depending on the country, 2 to 3 times the cases count during the 1^st^ “wave”) is entirely the effect of improved cases detection due to increased Testing Rate.

“Genuine Virus Prevalence” curves for the 1^st^ epidemic “wave” recorded a “Gompertz style” flattening and subsequent waning around 0.7% to 2% “depending on the country. Obviously these values are per-country averages and it is possible that at individual epidemic-outbreak localities the true virus prevalence might have been considerably higher. Nevertheless, it appears that epidemic curve flattening and waning happened at relatively modest levels of COVID-19 infection spread, either naturally or facilitated by the government measures that restricted social interaction.

## 5. Conclusions

This paper studied records of PCR Testing Rate and Test Positivity for nine European countries within the same geographic realm and recording similar trends in virus prevalence, and developed a method to quantify and remove the biasing effects of different Testing Rate (over time, or from one country to another) on Test Positivity. Bias-corrected records of Test Positivity called “genuine Virus Prevalence” are considered a reflection of the degree of community spread of COVID-19 infections. For the nine European countries, estimates of “genuine Virus Prevalence” were around 0.5 to 2% during the peak of the 1^st^ COVID-19 epidemic “wave” and are approaching similar levels in the ongoing 2^nd^ epidemic “wave”. It hence appears that in the 1^st^ epidemic “wave”, curve flattening and waning happened at relatively modest levels of COVID-19 infection spread, either naturally or facilitated by the government measures that restricted social interaction.

Results validation by comparison of “genuine Virus Prevalence” against Seroprevalence data for the studied countries gives a favourable outcome as differences between the two datasets are relatively small and the observed correlation significant. “Genuine Infection Fatality Rate” IFR, computed by pairing “genuine Virus Prevalence” with Excess Deaths data, also shows a reasonable match with closed-community model IFR. Finally, computed estimates of “genuine Virus Prevalence” for Slovakia give an acceptable match with the results of recent community mass testing in the country. Based on these validations, the method of computing “genuine Virus Prevalence” appears robust and may be deployable beyond just the initial dataset of countries.

## Supporting information

Reporting Standards checklist

## Data Availability

All datasets used for supporting the conclusions of this article are publically available from public data repositories as specified in the Methods section. Software used for statistical analysis is Microsoft Excel.

https://www.ecdc.europa.eu/en/covid-19/situation-updates-covid-19/covid-19-data/weekly

http://data.un.org/Data.aspx?d=POP&f=tableCode%3A22

## Ethics and Consent

Not applicable.

## Acknowledgements

The author would like to thank Dr DM Navaratnam from United Lincolnshire Hospital Trust, UK, Dr Malinee Thambyayah from Pantai Hospital Kuala Lumpur and Robert Everts from Almere, Netherlands, for their helpful review of this analysis and of earlier versions of the manuscript.

## Author Contributors

Single author.

